# A Review of Suicide-Related Risks through Propensity Score Methods using the Maryland Suicide Data Warehouse

**DOI:** 10.64898/2026.10.19.26364526

**Authors:** C Kitchen, JM Kush, A Zirikly, A Belouali, P Nestadt, HC Wilcox, H Kharrazi

## Abstract

**Objective:** A diversity of approaches and cohorts have been published in research literature for suicide risk estimation, leading to some uncertainty around which factors and conditions are most strongly contributing to risk. Methods exist for weighting estimates by propensity score, removing simplifying the interpretation of model effects. Few studies have explored the use of these tools to assess when the effect of certain factors is uncertain or inconsistent.

**Method:** A mix of 1,001,737 suicide decedents and living patients were evaluated for suicide risk, relying on a subset of commonly identified factors: demographics, psychiatric conditions, substance use disorders, and social needs. Care utilization patterns were then assessed controlling for these factors in a hospital care setting. Four methods were considered for controlling multivariate confounding of odds predicting death by suicide: multivariate logistic regression, 1-to-1 sample matching, inverse propensity weighting (IPTW), and entropy balancing (EBAL).

**Results:** Test statistics showed suicide decedents were much more likely to have various psychiatric and substance use diagnoses, social needs and more likely to heightened care utilization across sampling intervals. Greater utilization was found close to date of death. Effect sizes after IPTW and EBAL weighting were also generally greater than would have been documented by regression alone.

**Conclusions:** Propensity weighting is a useful alternative to exploring nuanced effects, especially when the direction and strength of an association differ from traditional statistics. By weighting, the overall imbalance of cases remains consistent and effect terms might reflect more nuanced associations than regression or sample matching alone.

## INTRODUCTION

Suicide risk analysis is often mired in methodological complexity, putting precise estimates of effects out of reach even with large sample sizes, rich sets of predictive features, and advanced modeling techniques. [1–4] Few significant predictors of suicide death are specific enough to enable targeted clinical decision support [2] or differentiate risks associated with self-harm without suicide intent. [5–7] Furthermore, the observational nature of research involving suicide death as an outcome prohibits most direct conclusions about such associations. [8–9] Nonetheless, clinicians infer that most known risk and protective factors for suicide are causal in nature since means of safely testing these effects in practice are lacking. [10]

Direct-observational and longitudinal research have been instrumental in identifying history of treatment, self-harm, bipolar or major depression, psychotic disorders, impulsivity, social factors, substance use, and physical illnesses as some of the strongest, most direct associations with suicide. [2] The mechanisms for each are at least partially well understood and reinforced by several meta-analytical studies. [1,2] Suicide risk, however, tends to be evaluated by patient reported symptoms and electronic health records (EHRs) rather than actual death. [1] These associations derived from observational research are not sufficient evidence for researchers to assert separate risks as causes of suicide death. Instead, the conventional view is that risks are additive, and suicide is simultaneously affected by multiple factors. [10]

Researchers have suggested that the effects of suicide predictors should be measured by propensity score weighting and matched sampling to overcome the limitations of observational designs. [9–10] Specifically, this can be summarized as transforming the natural incidence of exposure to risk factors simulating a randomized clinical trial, where direct relationships to outcomes can be quantified after controlling confounding variables. [8–10] This differs from the traditional mean-driven associationism approach of regression modeling, where observations are presumed to be independent [6], and non-collinearity is known to be faulty in practice. [4] In contrast, causal inference by propensity score weighting is one method for evaluating direct exposure effects, which has been deployed with some success in suicide risk assessment research. [8–10]

Prior research has shed light on several counterintuitive associations between known health risks and suicide death. For example, there are conflicting published findings on the role social determinants of health (SDoH) factors have in suicide risk. [11–15] Some known risk factors for other, unrelated health outcomes may appear to be negatively associated with suicide risk (e.g., legal involvement, homelessness, drug and alcohol use), though this is frequently explained by issues surrounding sample bias. [16–18] Others have tried to interpret these effects as more state and inequity-dependent phenomena, making them only a risk under certain conditions or result from drastic socio-economic change. [13,19] Though prior research has established some nuance to commonly identified risk factors, several clinical associations appear quite robust. Among them are mood, anxiety and psychotic disorder diagnoses with or without substance use disorders. [1,2,20,21] The availability, type and utilization of health care services over time is also known to be related to suicide, with many deaths among psychiatric patients occurring within just a month of hospital discharge. [22–25]

Current suicide risk analysis research might benefit from exploring the value of propensity score and matching methods to quantify exposure effects using large population-level suicide outcome data. [26] To address this gap, this study compares propensity weighting methods to quantify exposure effects across variable healthcare encounter time windows using the Maryland Suicide Data Warehouse (MSDW). [27] Lessons learned from this analysis have implications for developing state-based models for predictive modeling and the interpretation of significant risk factors of suicide. [6] Additionally, this study could reveal meaningful opportunities to develop new partnerships or enhance care coordination for specific types of providers across different time windows after controlling for extraneous confounding variables.

## METHODS

### Data and Participants

The MSDW has been developed for large-scale observational research on suicide and suicide-related outcomes in the State of Maryland. [27] The MSDW is a statewide repository of decedent information, as recorded through the State of Maryland Office of the Chief Medical Examiner (OCME). [28] The Chesapeake Regional Information System for our Patients (CRISP) is a regional health information exchange responsible for providing patient linkages across multiple data sources. [29] The MSDW includes patient records from multiple health systems and repositories, but the most relevant for this work are hospital discharge records from the Health Services Cost Review Commission (HSCRC) and encounters from the Johns Hopkins Medical Institution (JHMI). [30,31]

This project was reviewed and approved by the Johns Hopkins Bloomberg School of Public Health IRB (#00009022) and Maryland Department of Health IRB (protocol # 19-16). All data providers have approved a separate data use agreement (DUA) in accordance to our prior review and deidentified data were transmitted directly to the study team for secure storage and retrieval. Due to the nature of deidentified retrospective records, it was not feasible to obtain patient consent and thus was waived by the corresponding review boards.

Records from the HSCRC are administrative claims for hospital-based care services and include demographic, diagnostic and procedure coded events, captured at each health encounter and cover all visits for patients insured and uninsured. Similarly, the JHMI electronic health records provide encounter level records, and it is known that the confluence of claims and EHR derived clinical markers can address problems with data quality arising from either source used separately for prediction. [32,33]

A random sample of one million living control patients out of 6,554,990 total were identified among HSCRC data, each having at least one encounter between 2016 and 2020. A small number of patients (∼130) were found to have missing values for age or sex and subsequently discarded (N=999,870). There were an additional 1,867 decedents who had died by suicide in the same years, based on OCME determination, and had one or more encounters in either HSCRC or JHMI data. These decedents with clinical records in MSDW account for approximately 60.3% of all suicide decedents identified by OCME for this timeframe. [4,6]

### Variable Types

A subset of relevant clinical features was identified for bivariate, causal interpretation after accounting for the confounding effects of other features. [6] There were four categories of clinical features that could have been observed within the twelve months preceding suicide death. An additional six reflect encounter types within a variable time window. For controls, the index date is determined as a date selected at random during the observation period.

Clinical features were broadly defined as demographics, psychiatric diagnoses, substance use disorders, and social needs. [6] Demographic features included items reported at patient level, including patient sex, age at index date, and patient reported racial characteristics. Diagnostic features were limited to psychiatric diagnoses relevant to heightened suicide risk. [2,6] These diagnoses included the condition groupings of depressive disorders, bipolar disorder, psychotic disorders, anxiety disorder and post-traumatic stress, as defined in the Clinical Classifications Software Refined (CCSR). [34] Additional markers for alcohol, cannabis, opiate, stimulant and polysubstance use disorders were developed using the same CCSR definitions. [35] Social needs markers were limited to ICD-10-CM codes that have been curated by the SIREN workgroup and defined either as any non-specific social need, or having some selected relevance to health outcomes, which include housing instability, food insecurity, financial or transportation issues. [36] These four selected social needs have been the subject of study in our prior work. [14, 15, 37–39] Combined, the study included 17 clinical features in the analysis, with 12 exposure variables tested among diagnostic, substance use and social needs markers.

A secondary analysis was performed using type of encounter as a predictor for suicide death, to illustrate a possible change in predictive value after propensity weighting and against a gradient of time leading up to an index date. For living patients, this date was chosen randomly across the span of 2016-2020. The index date was the date of death for suicide decedents. Encounters were grouped in terms of point of care (e.g., inpatient, outpatient, emergency department) or by clinical intensity of care, including psychiatric hospitalization, having six or more encounters of any type or any psychiatric, group or family counseling services. The threshold of having at least 6 care encounters was decided from two considerations: because prior research in the frequency and timing of clinical encounters preceding suicide indicate roughly 7 – 12 encounters within a year more than doubled relative risk, and because the proportion of patients having 6 or more within a year was roughly 5 times greater among decedents in our own sample. [40] Raising this threshold of encounters for any time interval would have the negative effect of excluding too many patients to be differentially meaningful as an effect, especially with smaller observation intervals. There were 6 total encounter types explored across five time windows (7days, 1mos, 3mos, 6mos, 12mos), yielding a total of 30 exposure features. The 12mos time window was used for the descriptive summary statistics.

### Propensity Weighting, Matching and Causal Inference

Causal inference analysis makes use of sample weighting and partitioning around an exposure variable: the independent variable of interest for each tested association. Each observation of the exposure is weighted according to a matrix of covariates that are imbalanced between exposure groups. These weights put observations, in either the exposure group of the independent variable, on roughly a balanced footing for the confounding variables and enable a fairer evaluation of the exposure effect on the dependent variable. A vector of weights is then applied to a generalized linear model fit of each independent variables vs. dependent variable (IV-DV) interaction. [41]

Since each exposure is tested separately, it is possible to consider all other features as confounding variables that require balancing. [41] This ignores the real possibility of multiple observations being conditionally related to one another in at least two diagnostic categories, substance use disorders or social needs (many-to-many associations). For this reason, clinical features of the same type were omitted from the list of covariates used in each test (serial one-to-many associations). For example, when evaluating the effect of depressive disorders on the odds of suicide death in 12mos, bipolar disorder is not listed as a covariate, but alcohol use disorder is.

Propensity weighting and matched sampling through propensity weights make use of the average effect of the exposure group estimand (a.k.a., average treatment effect on treated individuals; ATT). [41] This meant weighting/matching was conducted with the exposure group as the reference for each comparison. Imbalance in covariate estimates was defined as at least a 10% difference between exposure groups on at least one covariate. The proportion of covariates imbalanced after weighting was monitored for issues in calibration and sampling. Following initial tests, effective sample size (ESS) was monitored to ensure no extreme imbalance between exposure groups was observed across covariates. [41] In a small number of exposures (i.e., depressive disorders, polysubstance use, and select social needs), the exposure group had a modestly larger ESS than the control group in the inverse probability of treatment weighting (IPTW). It was then decided that weights for IPTW and entropy balancing (EBAL) methods required trimming at the 99^th^ percentile to prevent loss due to incomparable observations. These were rare instances of extreme clinical presentation [4] resulting in an imbalance of ESS.

Effect sizes for each IV were translated into odds bounded by 95% confidence intervals for plotting and ease of interpretation across each balancing method. Results for four regression-based approaches were assessed: (1) the logistic model coefficient for the exposure variable, with associated covariates as confounds, (2) the model association for a 1-to-1 matched sample using a nearest neighbor optimization strategy on confounding effects, (3) the IV-DV model association after IPTW weighting, and (4) the IV-DV model association after EBAL weighting. Covariate balancing, regression modeling and propensity score weighting was achieved using the *survey* and *WeightIt* packages of the R programming language. [42,43] Sample matching by nearest neighbor distance was conducted using the *MatchIt* R package. [44]

## RESULTS

Patient level characteristics differed significantly between suicide decedents and living controls (**Table 1**). Suicide decedents were found to be 10 years older on average (t = 23.8; p < 0.001) and were 75% male, compared to 46.7% among living controls (χ2 = 599.8; p < 0.001). Decedents were also significantly more likely to identify as Caucasian (χ2 = 515.3; p < 0.001) and be divorced, widowed or separated (χ2 = 187.5; p < 0.001).

**TABLE 1.** Clinical and demographic characteristics for suicide decedents and hospital patients.

| Covariate | Living | Suicide | Chi Squared | Naïve Odds |
| --- | --- | --- | --- | --- |
| Total Sample | 999,870 | 1,867 |  |  |
| Patient Level Features |  |  |  |  |
| Average Age in Years | 39.0 (SD=24.7) | 49.9 (SD=19.7) |  |  |
| Female | 532,864 (53.3%) | 466 (25%) | 599.8* |  |
| Male | 467,006 (46.7%) | 1,401 (75%) |  |  |
| African American | 271,218 (27.1%) | 284 (15.2%) | 515.3* |  |
| Caucasian | 486,742 (48.7%) | 1,397 (74.8%) |  |  |
| Hispanic/Latino | 90,031 (9%) | 73 (3.9%) |  |  |
| Other or Unknown Race | 151,879 (15.2%) | 113 (6.1%) |  |  |
| Divorced, Separated or Widowed | 96,962 (9.7%) | 356 (19.1%) | 187.5* |  |
| Married | 343,796 (34.4%) | 595 (31.9%) |  |  |
| Single, or Unknown Marital Status | 559,112 (55.9%) | 916 (49.1%) |  |  |
| Medicaid Enrollment | 65,004 (6.5%) | 214 (11.5%) | 788.7* |  |
| Medicare Enrollment | 156,477 (15.6%) | 629 (33.7%) |  |  |
| Other or Unknown Payer | 639,899 (64%) | 633 (33.9%) |  |  |
| Private Insurance | 138,490 (13.9%) | 391 (20.9%) |  |  |
| Features Occurring on Clinical Encounters within 12mos of Index Date |  |  |  |  |
| Depressive Disorder | 27,574 (2.8%) | 497 (26.6%) | 3887.1* | 12.8 |
| Anxiety Disorder | 17,515 (1.8%) | 431 (23.1%) | 4808.4* | 16.8 |
| PTSD | 3,659 (0.4%) | 142 (7.6%) | 2564.9* | 22.4 |
| Bipolar Disorder | 3,867 (0.4%) | 173 (9.3%) | 3635.8* | 26.3 |
| Psychotic Disorder | 2,689 (0.3%) | 104 (5.6%) | 1864.7* | 21.9 |
| Alcohol Use Disorder | 8,442 (0.8%) | 216 (11.6%) | 2489.2* | 15.4 |
| Opiate Use Disorder | 4,048 (0.4%) | 111 (5.9%) | 1370.2* | 15.6 |
| Stimulant Use Disorder | 2,343 (0.2%) | 73 (3.9%) | 1031.2* | 17.3 |
| Polysubstance Use Disorder | 3,667 (0.4%) | 124 (6.6%) | 1929.6* | 19.3 |
| Cannabis Use Disorder | 6,212 (0.6%) | 112 (6%) | 850.5* | 10.2 |
| Selected Social Needs (x4) | 1,823 (0.2%) | 51 (2.7%) | 635* | 15.4 |
| General Social Needs (x1) | 873 (0.1%) | 27 (1.4%) | 368.4* | 16.8 |
| Encounter Types by Point of Care within 12mos of Index Date |  |  |  |  |
| 1+ Outpatient Visit | 343,977 (34.4%) | 1,257 (67.3%) | 893* | 3.9 |
| 1+ Inpatient Stay | 83,342 (8.3%) | 590 (31.6%) | 1311* | 5.1 |
| 1+ Emergency Department Visit | 50,097 (5.0%) | 586 (31.4%) | 2693.6* | 8.7 |
| Any Psychiatric, Group or Family Counseling | 5,280 (0.5%) | 63 (3.4%) | 279.2* | 6.6 |
| Psychiatric Hospitalization | 1,711 (0.2%) | 128 (6.9%) | 4508* | 42.9 |
| Greater than or Equal to 6 Encounters | 54,600 (5.5%) | 526 (28.2%) | 1844.3* | 6.8 |

\* Significant at 0.001

Covariates and exposure features were assessed along with naïve odds of suicide death as an outcome for each exposure, without considering any confounding effect from other variables (**Table 1**). All exposure variables were found to have significant and elevated test statistics on a simple 2 x 2 chi squared test of independence. Odds of suicide death were several times greater for each exposure, and most noteworthy for bipolar disorder (OR = 26.3), post-traumatic stress disorder (OR = 22.4), psychotic disorders (OR = 21.9), and polysubstance use disorder (OR = 19.3).

Features derived from encounter types and tested along a moving time window were generally less significantly associated with suicide death (**Table 1**). Each tested association remained significant on a chi squared test of independence. Without controlling for confounding features, suicide death was found to be roughly 4 to 9 times more likely for most types of encounters within 12 months of the index date. For psychiatric hospitalization, the odds were 42.9 times greater (**Table 1**).

Adjusted effects of exposure variables were converted to odds ratios for ease of interpretation across each type of estimate (**Figure 1**). The propensity weighted estimates were larger in size and variability than either binary logistic regression or matched sample. The matched sample estimates were found to be the smallest, though most remained statistically significant (**Table 2**). Among psychiatric conditions, the propensity weighted ORs were significantly greater than the point estimate of the logistic regression for psychotic disorders (IPTW’s OR = 7.97). The same scores were marginally greater for bipolar disorders and post-traumatic stress (OR = 9.32; OR = 8.70, respectively), but lower or mixed for depressive and anxiety disorders (OR = 4.76; OR = 6.61) and not significantly different from model estimates. Each of the matched sample effects was somewhat smaller in size, with point estimates for psychotic and anxiety disorders being the two highest conditions related with suicide death (OR = 3.96; OR = 3.60) (**Table 2**).

**FIGURE 1.**
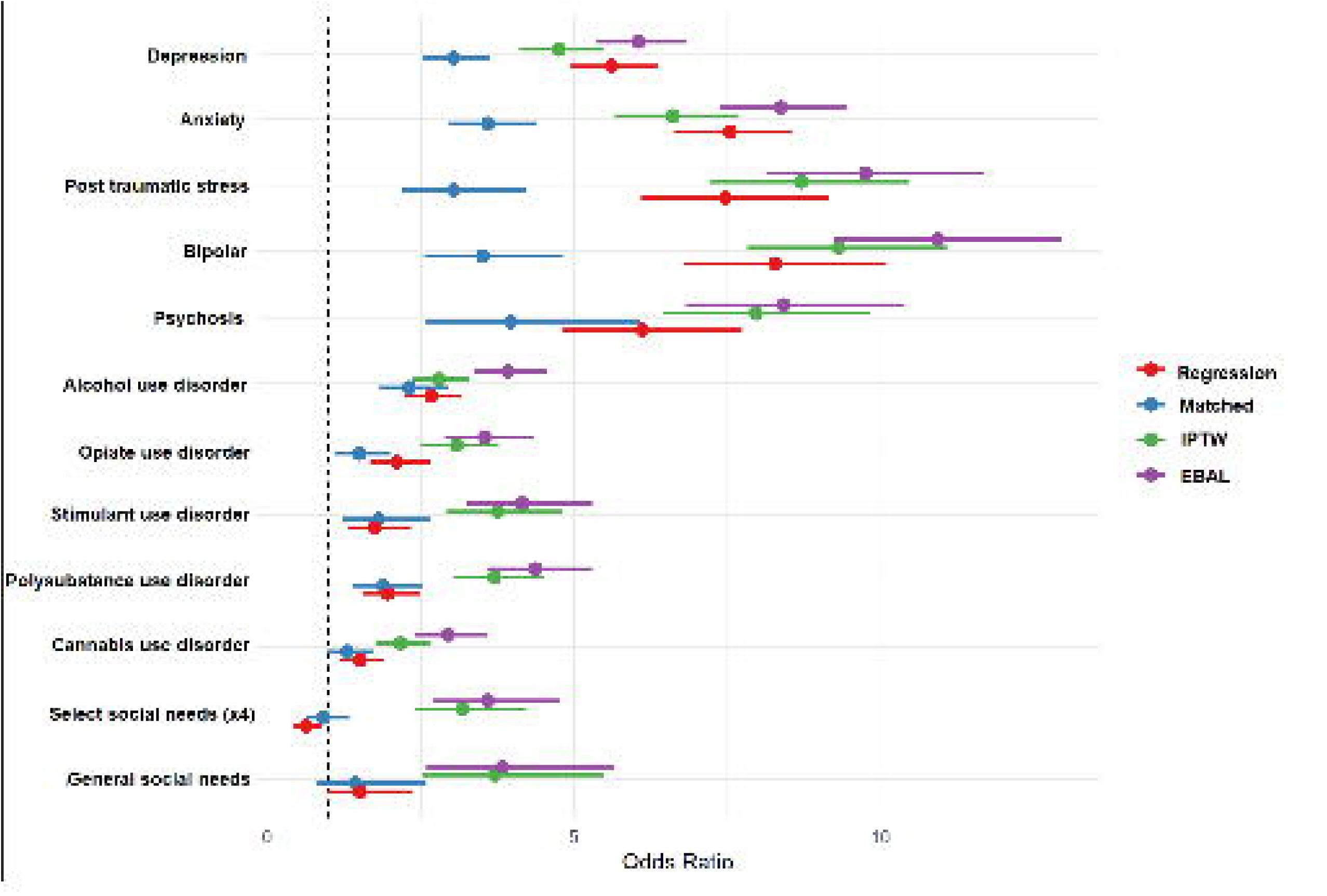
Individual 12 month effects expressed as odds ratios of exposure variables across each type of weighting method.

**TABLE 2.** **O**dds ratios from clinical exposure variables across multivariate regression and different weighting methods: inverse pro**bability** treatment weighting (IPTW), entropy balancing (EBAL) and sample matching.

| Exposure Variable | Logistic Regression | Matched Sample | IPTW | EBAL |
| --- | --- | --- | --- | --- |
| Depressive Disorder | 5.61 (4.95:6.36) | 3.03 (2.54:3.61) | 4.76 (4.11:5.5) | 6.05 (5.36:6.83) |
| Anxiety Disorder | 7.54 (6.64:8.57) | 3.6 (2.94:4.4) | 6.61 (5.68:7.69) | 8.36 (7.4:9.45) |
| PTSD | 7.47 (6.09:9.16) | 3.03 (2.18:4.22) | 8.7 (7.24:10.47) | 9.75 (8.13:11.7) |
| Bipolar Disorder | 8.28 (6.8:10.09) | 3.5 (2.55:4.8) | 9.32 (7.84:11.08) | 10.94 (9.25:12.95) |
| Psychotic Disorder | 6.1 (4.81:7.74) | 3.96 (2.59:6.07) | 7.97 (6.45:9.84) | 8.41 (6.82:10.38) |
| Alcohol Use Disorder | 2.66 (2.25:3.15) | 2.31 (1.81:2.94) | 2.79 (2.37:3.27) | 3.91 (3.36:4.56) |
| Opiate Use Disorder | 2.1 (1.67:2.65) | 1.49 (1.11:2.01) | 3.08 (2.51:3.78) | 3.54 (2.89:4.34) |
| Stimulant Use Disorder | 1.74 (1.3:2.34) | 1.8 (1.23:2.66) | 3.75 (2.93:4.8) | 4.15 (3.25:5.3) |
| Polysubstance Use Disorder | 1.96 (1.55:2.48) | 1.88 (1.39:2.54) | 3.7 (3.04:4.51) | 4.36 (3.59:5.3) |
| Cannabis Use Disorder | 1.5 (1.19:1.89) | 1.29 (0.97:1.71) | 2.17 (1.76:2.66) | 2.94 (2.41:3.6) |
| Select Social Needs (x4) | 0.63 (0.44:0.89) | 0.91 (0.62:1.33) | 3.18 (2.39:4.24) | 3.59 (2.69:4.77) |
| General Social Needs (x1) | 1.51 (0.97:2.35) | 1.43 (0.79:2.6) | 3.71 (2.51:5.47) | 3.83 (2.59:5.65) |

Other independent variables for substance use disorders and social needs within 12 months of death yielded considerably smaller effects (**Table 2**). Propensity weighted ORs of IPTW were significant and significantly greater than those from the logistic regression in opiate use disorders (OR = 3.08), stimulant use disorders (OR = 3.75), polysubstance use disorders (OR = 3.70) and cannabis use disorder (OR = 2.17), but not alcohol use disorder (OR = 2.79). Select social needs and any coded social need markers were both significant (OR = 3.18; OR = 3.71) and much larger than either the matched sample (OR = 0.91; OR = 1.43) or logistic regression estimates (OR = 0.63; OR = 1.51).

A ratio of exposure-to-control size was inspected as a check on ESS validity for each estimate (**Figure 2**). This ratio was seen to exceed an acceptable threshold for drawing conclusions in only three tests: the IPTW estimates of twelve-month exposures to depressive disorders, polysubstance use and select social needs. The effects produced similar results with acceptable ESS as a contrast to the EBAL method of weighting (**Table 2**).

**FIGURE 2.**
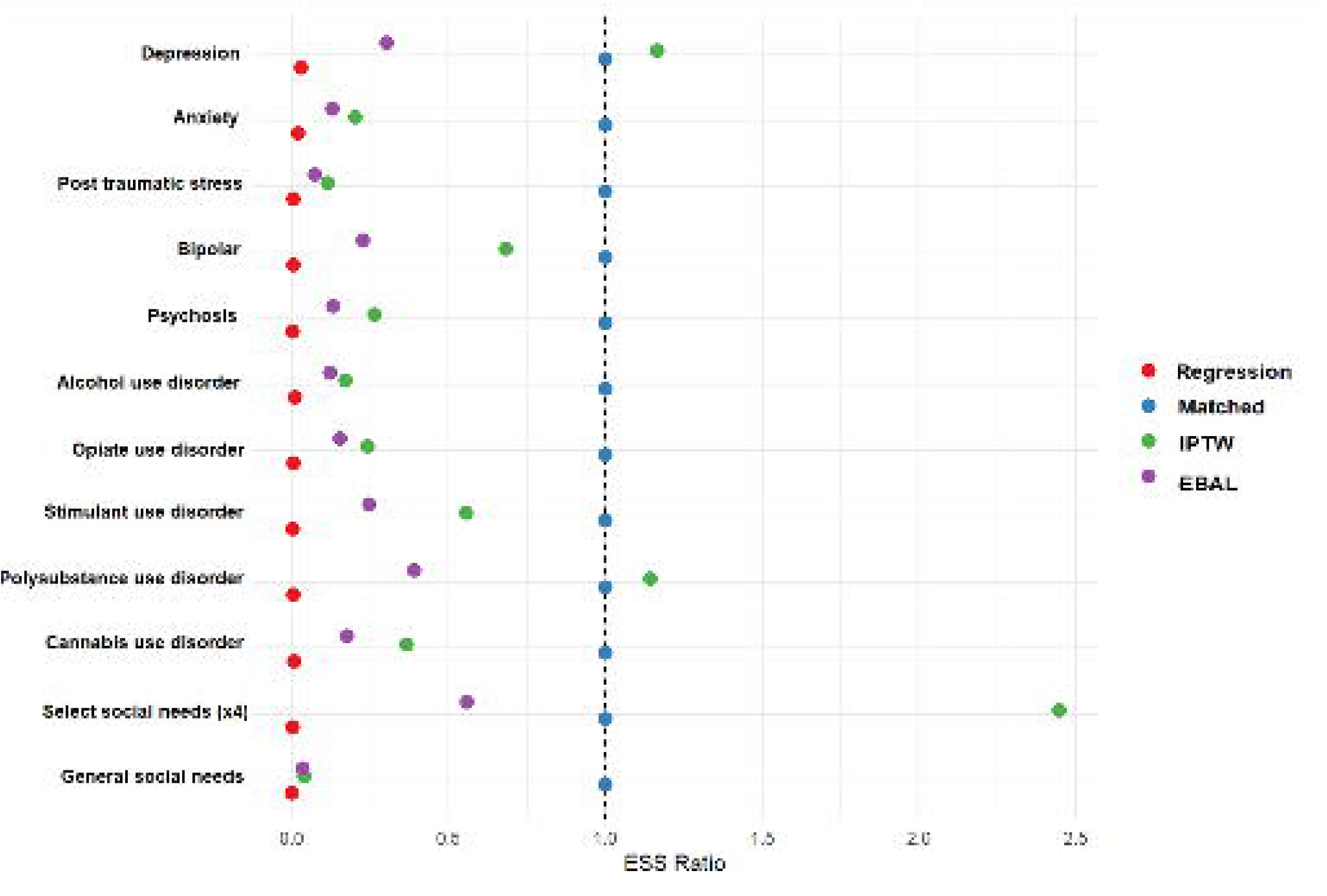
Effective sample size (ESS), ratio of exposure-to-control for each estimate.

The encounter type and time horizon analysis yielded similar results for differential performance of weighting methods. Samples matched 1-to-1 on demographics, clinical features, and social needs yielded the lowest overall estimates, but generally were statistically significant (**Table 3**). The presence of an outpatient encounter within time windows leading up to the date of death had little-to-no significant IPTW relation to suicide as an outcome (OR = 0.99; 1.03; 1.04; 1.05; 1.19). Emergency department encounters were significant and most related to suicide death at 12 months and within 7 days to death across all estimate types. All cause hospitalization had a similarly increasing relationship with suicide death across time horizons, but psychiatric hospitalizations were consistently associated with smaller odds, as the index date became closer (Supplemental Figure 1). Having six or more encounters of any type was positively and strongly associated with suicide death at all time horizons and methods of estimating effects. This was progressively stronger with temporal proximity to death date, especially at 7 days. As many as 98 times as many decedents were found to have 6+ encounters within 7 days of index, compared to living controls, when viewed as a predictive ratio between either group (Supplemental Table 1). Counseling sessions, emergency room visitation, and all cause hospitalization followed this progressive trajectory, which underscores the frequency, timing and type of encounters all contributed to our ability to assess risk. Propensity weighting yielded suitable ESS on all but outpatient care service estimates and only at longer intervals at 3 months or more (Supplemental Figure 2).

**TABLE 3.**
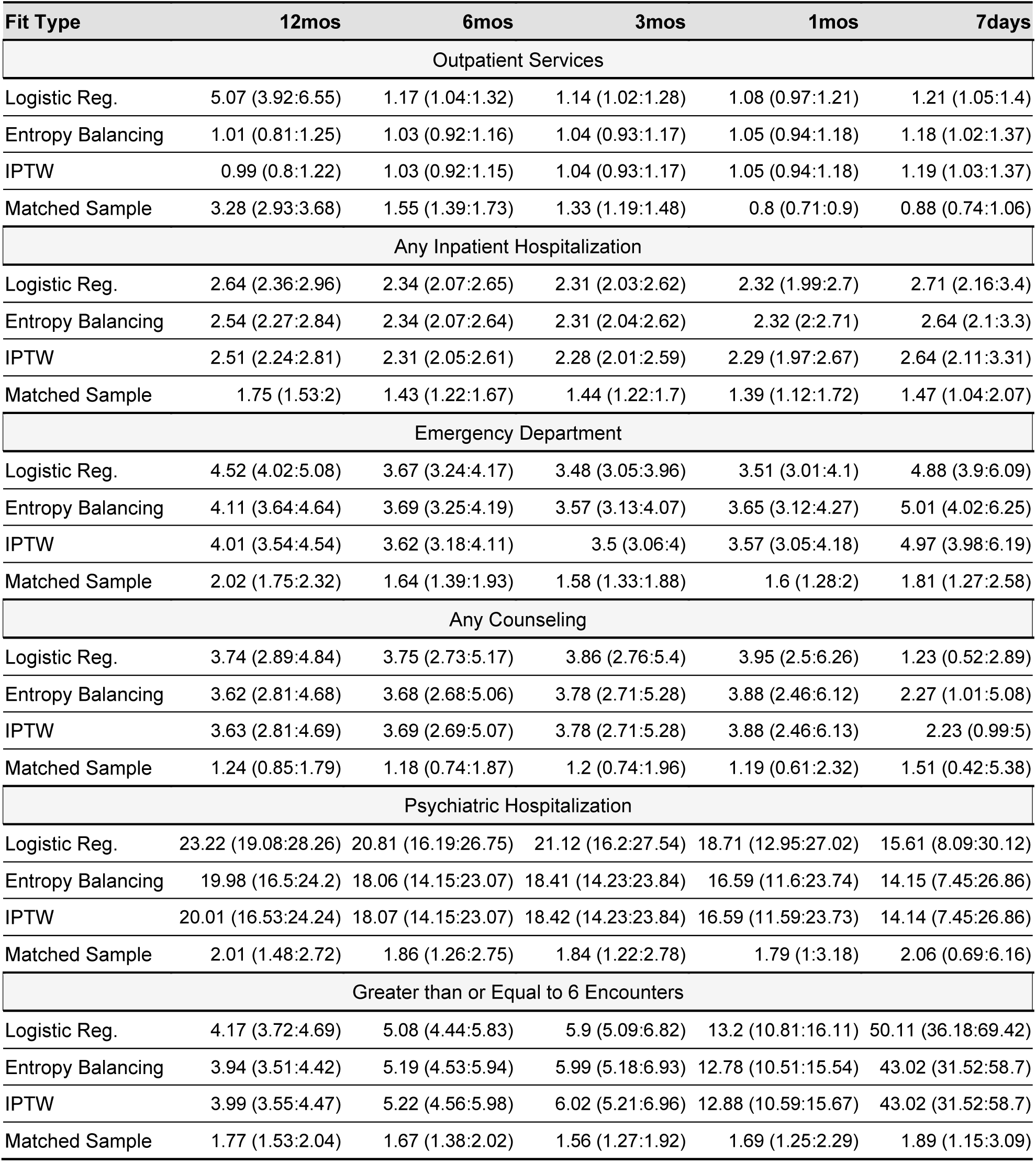
**O**dds ratios from time horizon and care utilization across multivariate regression and different weighting methods: inverse probability treatment weighting (IPTW), entropy balancing (EBAL) and sample matching.

| Fit Type | 12mos | 6mos | 3mos | 1mos | 7days |
| --- | --- | --- | --- | --- | --- |
| Outpatient Services |  |  |  |  |  |
| Logistic Reg. | 5.07 (3.92:6.55) | 1.17 (1.04:1.32) | 1.14 (1.02:1.28) | 1.08 (0.97:1.21) | 1.21 (1.05:1.4) |
| Entropy Balancing | 1.01 (0.81:1.25) | 1.03 (0.92:1.16) | 1.04 (0.93:1.17) | 1.05 (0.94:1.18) | 1.18 (1.02:1.37) |
| IPTW | 0.99 (0.8:1.22) | 1.03 (0.92:1.15) | 1.04 (0.93:1.17) | 1.05 (0.94:1.18) | 1.19 (1.03:1.37) |
| Matched Sample | 3.28 (2.93:3.68) | 1.55 (1.39:1.73) | 1.33 (1.19:1.48) | 0.8 (0.71:0.9) | 0.88 (0.74:1.06) |
| Any Inpatient Hospitalization |  |  |  |  |  |
| Logistic Reg. | 2.64 (2.36:2.96) | 2.34 (2.07:2.65) | 2.31 (2.03:2.62) | 2.32 (1.99:2.7) | 2.71 (2.16:3.4) |
| Entropy Balancing | 2.54 (2.27:2.84) | 2.34 (2.07:2.64) | 2.31 (2.04:2.62) | 2.32 (2:2.71) | 2.64 (2.1:3.3) |
| IPTW | 2.51 (2.24:2.81) | 2.31 (2.05:2.61) | 2.28 (2.01:2.59) | 2.29 (1.97:2.67) | 2.64 (2.11:3.31) |
| Matched Sample | 1.75 (1.53:2) | 1.43 (1.22:1.67) | 1.44 (1.22:1.7) | 1.39 (1.12:1.72) | 1.47 (1.04:2.07) |
| Emergency Department |  |  |  |  |  |
| Logistic Reg. | 4.52 (4.02:5.08) | 3.67 (3.24:4.17) | 3.48 (3.05:3.96) | 3.51 (3.01:4.1) | 4.88 (3.9:6.09) |
| Entropy Balancing | 4.11 (3.64:4.64) | 3.69 (3.25:4.19) | 3.57 (3.13:4.07) | 3.65 (3.12:4.27) | 5.01 (4.02:6.25) |
| IPTW | 4.01 (3.54:4.54) | 3.62 (3.18:4.11) | 3.5 (3.06:4) | 3.57 (3.05:4.18) | 4.97 (3.98:6.19) |
| Matched Sample | 2.02 (1.75:2.32) | 1.64 (1.39:1.93) | 1.58 (1.33:1.88) | 1.6 (1.28:2) | 1.81 (1.27:2.58) |
| Any Counseling |  |  |  |  |  |
| Logistic Reg. | 3.74 (2.89:4.84) | 3.75 (2.73:5.17) | 3.86 (2.76:5.4) | 3.95 (2.5:6.26) | 1.23 (0.52:2.89) |
| Entropy Balancing | 3.62 (2.81:4.68) | 3.68 (2.68:5.06) | 3.78 (2.71:5.28) | 3.88 (2.46:6.12) | 2.27 (1.01:5.08) |
| IPTW | 3.63 (2.81:4.69) | 3.69 (2.69:5.07) | 3.78 (2.71:5.28) | 3.88 (2.46:6.13) | 2.23 (0.99:5) |
| Matched Sample | 1.24 (0.85:1.79) | 1.18 (0.74:1.87) | 1.2 (0.74:1.96) | 1.19 (0.61:2.32) | 1.51 (0.42:5.38) |
| Psychiatric Hospitalization |  |  |  |  |  |
| Logistic Reg. | 23.22 (19.08:28.26) | 20.81 (16.19:26.75) | 21.12 (16.2:27.54) | 18.71 (12.95:27.02) | 15.61 (8.09:30.12) |
| Entropy Balancing | 19.98 (16.5:24.2) | 18.06 (14.15:23.07) | 18.41 (14.23:23.84) | 16.59 (11.6:23.74) | 14.15 (7.45:26.86) |
| IPTW | 20.01 (16.53:24.24) | 18.07 (14.15:23.07) | 18.42 (14.23:23.84) | 16.59 (11.59:23.73) | 14.14 (7.45:26.86) |
| Matched Sample | 2.01 (1.48:2.72) | 1.86 (1.26:2.75) | 1.84 (1.22:2.78) | 1.79 (1:3.18) | 2.06 (0.69:6.16) |
| Greater than or Equal to 6 Encounters |  |  |  |  |  |
| Logistic Reg. | 4.17 (3.72:4.69) | 5.08 (4.44:5.83) | 5.9 (5.09:6.82) | 13.2 (10.81:16.11) | 50.11 (36.18:69.42) |
| Entropy Balancing | 3.94 (3.51:4.42) | 5.19 (4.53:5.94) | 5.99 (5.18:6.93) | 12.78 (10.51:15.54) | 43.02 (31.52:58.7) |
| IPTW | 3.99 (3.55:4.47) | 5.22 (4.56:5.98) | 6.02 (5.21:6.96) | 12.88 (10.59:15.67) | 43.02 (31.52:58.7) |
| Matched Sample | 1.77 (1.53:2.04) | 1.67 (1.38:2.02) | 1.56 (1.27:1.92) | 1.69 (1.25:2.29) | 1.89 (1.15:3.09) |

## DISCUSSION

Suicide risk analysis often uses multivariate methods to measure the effect of predictors on suicide outcomes. Such observational research relies on conditional mean effects to estimate extraneous variables. However, this approach becomes an issue when the direction or significance of an effect defies expectations or is subject to substantial collinearity. To address this challenge, this study compares the results of a traditional regression approach to results of different propensity matching methods in assessing the effect of known predictors of suicide. Using these methods, counterintuitive effects can be easily evaluated with confidence intervals using the same set of covariates in tandem with causal inference analysis.

Collectively, the findings of this study were consistent with prior research. Select predictors, similar to variables studied in the past, showed a significant and substantial association with suicide death, after controlling for multiple, potentially confounding covariates. Many of the most predictive diagnostic categories for suicidal thoughts and behaviors (STB) are at best weak predictors of suicide. In a recent meta analysis of literature, peak weighted OR of depressive and anxiety disorders were estimated around 2.45 and 1.79, respectively (wOR:∼1.6 for general psychopathology predicting suicide death). [2] Follow-up intervals for most studies in this meta analysis were on the order of several years, which authors hypothesized could have washed out any time-varying effects of acute stressors more reliably predicting STB. Our own 12-month effects for depressive and anxiety disorders, along with plausible between group bias from disparate care utilization likely resulted in somewhat larger effect sizes on average than what is seen elsewhere in literature. [2,24]

Controlling confounding effects may be the single best reason for employing these methods as demonstrated by the evaluation of naïve odds for each IV-DV association (**Table 1**), or through logistic models. When exposure variables are weighted, the effects look drastically different (**Table 2**). For example, initially the base logistic regression showed that select social needs are significantly associated with a reduction in odds of suicide death (OR = 0.63). However, after accounting for the same set of covariates, using IPTW and EBAL weights, the association between select social needs and suicide death appeared more consistent with expectations (OR

= 3.18 and 3.59, respectively). Corroborating this observation, we found the aforementioned meta analysis of pooled effects had a wOR of 2.20 (CI 1.32:3.67) for what authors describe as “low socioeconomic status.” [2]

Prior research has found time-varying factors as being highly predictive of risk, particularly discharge from psychiatric hospitalization. This risk seems more acute shortly after discharge, as it has been reported that incidence of suicide among recent discharges is several times higher than the US national average. [23,24] One study found approximately 0.28% of all discharged psychiatric patients died from suicide within 3 months but most decedents were not hospitalized just prior to death. [23] Still, we found psychiatric hospitalization was highly predictive of suicide death among all hospital patients at 12 months (OR: 23.22 CI: 19.08:28.26), but this logistic effect diminished as the observation window approached 7 days of index (OR: 14.15 CI:7.45:28.88), suggesting risk is not elevated upon psychiatric discharge. Overall care utilization, including outpatient face-to-face contacts, are reliably associated with suicide risk, with one Danish study reporting roughly 64.9% more decedents had outpatient care in the month prior to death (prevalence, 32.0%), compared to the general population. [22] Our study found an analogous 90.6% more decedents with outpatient care (prevalence, 26.3%) compared to living controls within 1 month of death. Direct comparison of effect sizes is problematic, as we have not identified research making use of hospital discharge records in precisely the same way to reflect general living control patients of a suicide risk modeling cohort. These results demonstrate that, for many who go on to die by suicide, there are repeated contacts with health services shortly before death. This is at least partially corroborated with literature that identifies significant differences in care service utilization for suicide decedents compared with non-deceased hospital patients, especially in the month prior to death. [22,23]

Though it may be uncommon to consider all types of care encounters as indicators of risk in the literature, it is undeniable that temporal effects are present. Patients susceptible to risk factors are frequently found to make contact at some level. Researchers evaluating suicide risk among drug users found health care encounters around 5 times more prevalent among decedents than living controls, matched by age, sex, marital status, education, alcohol use disorder and psychiatric condition. This difference shrank to being roughly equivocal between groups at 1 year. Our work found these temporal effects most pronounced in identifying those with 6 or more discrete hospital-based encounters within 7 days of death. Roughly 2.6% of suicide decedents met this threshold, while just 0.0003% of living hospital controls did, yielding a predictive ratio of 97.9 and logistic regression OR of 50.11 (CI 36.18:69.42). At 1 month, this ratio was much smaller, but still among the largest effect sizes observed in this study (OR: 13.20, CI 10.81:16.11). Though the alternatively estimated effects, using IPTW, EBAL and sample matching were smaller, a significant and substantial association was found consistently across time intervals, peaking at the 7 day window.

Every point of contact with a patient represents an opportunity to intervene or screen risk. Analysts might be served by propensity weighting techniques as a mechanism for not surreptitiously disregarding highly predictive features simply due to collinearity, as might occur in automated feature selection tasks, like least absolute shrinkage and selection operator (i.e., LASSO). Weighting might also better inform expectations of exposures in retrospective observational research, especially where simple matching might introduce unwanted artifacts to these estimates. Clinicians care to know which factors are contributing most to risk. This is a different kind of question than asking “by how much” and one that might best be answered with an ordinal ranking. It may not matter much for providers that the odds ratio estimate for bipolar disorders was significantly greater in logistic regression compared to matched sample estimates, it just matters for prediction and classification. For analysts, this means much needed transparency in which variables to use when fitting a predictive model, even if using nonparametric tools to circumvent issues of collinearity. To stakeholders, having a clearer idea of the real versus theoretical effect sizes is still important for managing care. To that end, both propensity weighting and matching techniques tended to alter the effects obtained through regression analysis.

Despite some usefulness of propensity matching methods, the study has several limitations. First, this study was limited to the population represented in the MSDW, hence any limitations represented in the underlying data could affect the results (e.g., population composition, data quality biases). [45] Second, MSDW captures hospital discharge claims, health information exchange data, and the medical examiner findings; however, it does not capture full EHR data and data, specifically clinical notes. Such clinical texts include additional diagnoses or social needs missing in structured data used in this study. [46,47] Third, the list of variables used for the analysis was a subset of available indicators selected based on prior studies, including our own research. Changing the underlying variables used as covariates may drastically change resulting effect estimates. It was not feasible or necessary to exhaustively test all potentially confounding attributes to reach our main findings. [6,17] Fourth, the methods proposed for the analysis were intended to make interpretation of effects easier, and thus fell short of being a formal analysis in either mechanisms of action or causation. The study relied on weighted GLM estimates of effects rather than separate estimand statistics. This approach was intended to generate individual odds ratios of coefficient estimates with confidence intervals that are more familiar for clinical researchers. [7,9]

Lastly, reliance on the ATT weights may be problematic, since the typical observational study does not perform non-random under-sampling of control subjects. This approach is only useful if it sufficiently mimics the gold standard of a randomized clinical trial. Understandably, the 1-to-1 matched sampling method yielded the most equivocal effect sizes and illustrates how control group undersampling might result in underestimation of effect sizes. [48] It is unclear whether other weighting strategies had a similarly distorting effect in the opposite direction.

## CONCLUSION

Suicide risk research is likely to continue exploring various analytical methods to understand nuanced interactions and effect size estimates of suicide outcome predictors. Using propensity score weights and matched samples could provide a useful solution to assess such predictors; however, as their use hinges on the appropriate identification of covariates the results should be interpreted cautiously. [7,9] Propensity weights have also been used with some success to pre-treat health data for the removal of algorithmic bias. Such applications may become a more trustworthy use case in the near future, especially where temporal proximity of clinical encounters is considered.

## Data Availability

Due to the nature of these data and explicit restriction from use other than for our specific protocol, per both conditions of IRB approval and DUA, these data are not available for secondary use.

## ACKNOWLEDGEMENTS

The authors of this work are grateful for the dedicated efforts of Elyse Lasser and Thomas Richards of the Center for Population Health IT. This work was possible because of their familiarity with managing multiple data agreements and databases, throughout the project.

## SUPPLEMENTAL

**TABLE S1.**
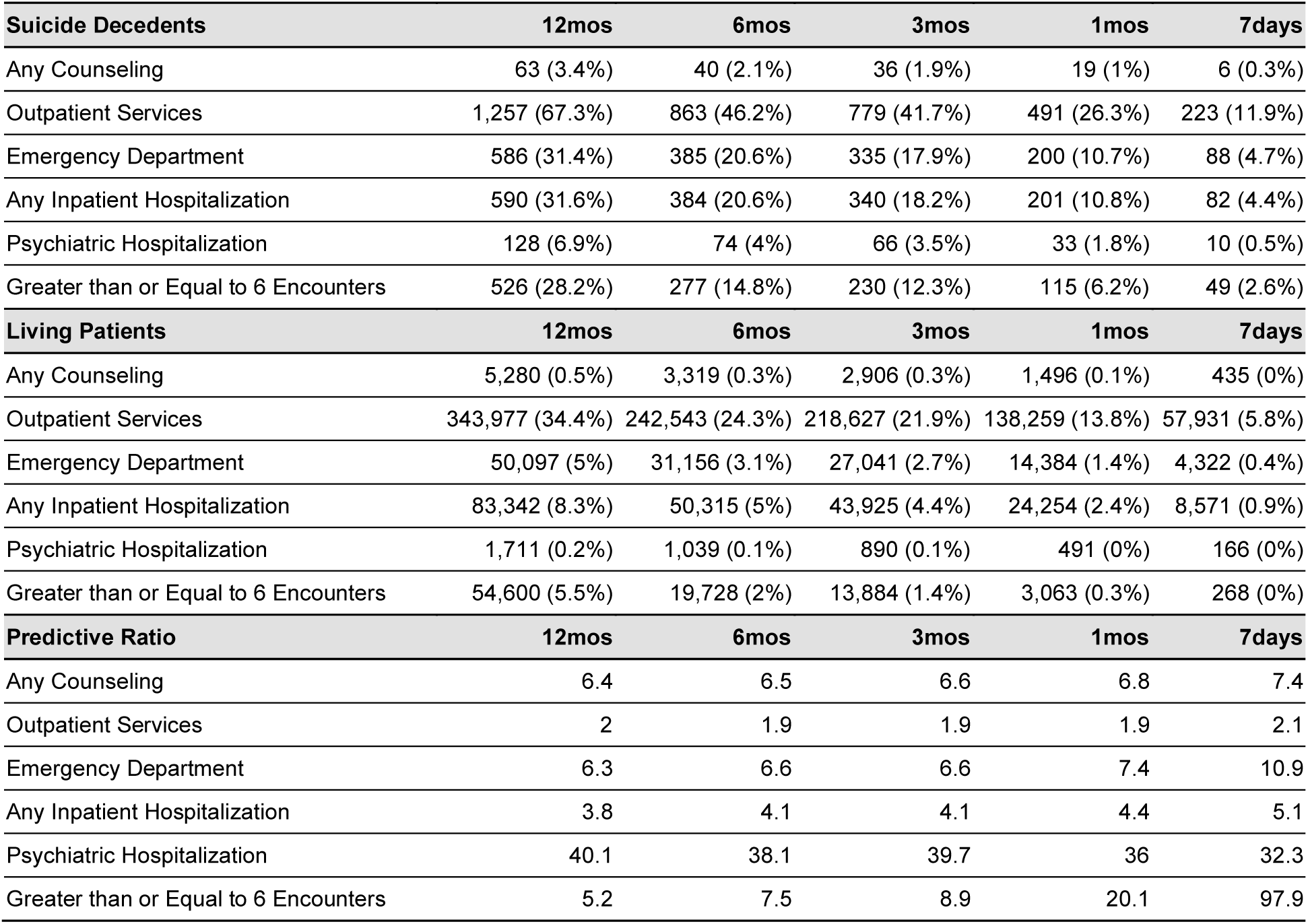
Counts of patients and corresponding predictive ratios across various time horizons and methods among suicide decedents and living control populations.

FIGURE S1. Effects of exposure variables as odds ratio across each type of weighting method and time horizon.

FIGURE S2. Effective sample size (ESS), ratio of exposure-to-control size for each estimate and time horizon

