## Supplemental for "A Review of Suicide-Related Risks through Propensity Score Methods using the Maryland Suicide Data Warehouse"

**APPENDIX**

**TABLE S1.** Counts of patients and corresponding predictive ratios across various time horizons and methods among suicide decedents and living control populations.

| **Suicide Decedents** | **12mos** | **6mos** | **3mos** | **1mos** | **7days** |
| --- | --- | --- | --- | --- | --- |
| Any Counseling | 63 (3.4%) | 40 (2.1%) | 36 (1.9%) | 19 (1%) | 6 (0.3%) |
| Outpatient Services | 1,257 (67.3%) | 863 (46.2%) | 779 (41.7%) | 491 (26.3%) | 223 (11.9%) |
| Emergency Department | 586 (31.4%) | 385 (20.6%) | 335 (17.9%) | 200 (10.7%) | 88 (4.7%) |
| Any Inpatient Hospitalization | 590 (31.6%) | 384 (20.6%) | 340 (18.2%) | 201 (10.8%) | 82 (4.4%) |
| Psychiatric Hospitalization | 128 (6.9%) | 74 (4%) | 66 (3.5%) | 33 (1.8%) | 10 (0.5%) |
| Greater than or Equal to 6 Encounters | 526 (28.2%) | 277 (14.8%) | 230 (12.3%) | 115 (6.2%) | 49 (2.6%) |
| **Living Patients** | **12mos** | **6mos** | **3mos** | **1mos** | **7days** |
| Any Counseling | 5,280 (0.5%) | 3,319 (0.3%) | 2,906 (0.3%) | 1,496 (0.1%) | 435 (0%) |
| Outpatient Services | 343,977 (34.4%) | 242,543 (24.3%) | 218,627 (21.9%) | 138,259 (13.8%) | 57,931 (5.8%) |
| Emergency Department | 50,097 (5%) | 31,156 (3.1%) | 27,041 (2.7%) | 14,384 (1.4%) | 4,322 (0.4%) |
| Any Inpatient Hospitalization | 83,342 (8.3%) | 50,315 (5%) | 43,925 (4.4%) | 24,254 (2.4%) | 8,571 (0.9%) |
| Psychiatric Hospitalization | 1,711 (0.2%) | 1,039 (0.1%) | 890 (0.1%) | 491 (0%) | 166 (0%) |
| Greater than or Equal to 6 Encounters | 54,600 (5.5%) | 19,728 (2%) | 13,884 (1.4%) | 3,063 (0.3%) | 268 (0%) |
| **Predictive Ratio** | **12mos** | **6mos** | **3mos** | **1mos** | **7days** |
| Any Counseling | 6.4 | 6.5 | 6.6 | 6.8 | 7.4 |
| Outpatient Services | 2 | 1.9 | 1.9 | 1.9 | 2.1 |
| Emergency Department | 6.3 | 6.6 | 6.6 | 7.4 | 10.9 |
| Any Inpatient Hospitalization | 3.8 | 4.1 | 4.1 | 4.4 | 5.1 |
| Psychiatric Hospitalization | 40.1 | 38.1 | 39.7 | 36 | 32.3 |
| Greater than or Equal to 6 Encounters | 5.2 | 7.5 | 8.9 | 20.1 | 97.9 |

**FIGURE S1.** Effects of exposure variables as odds ratio across each type of weighting method and time horizon.

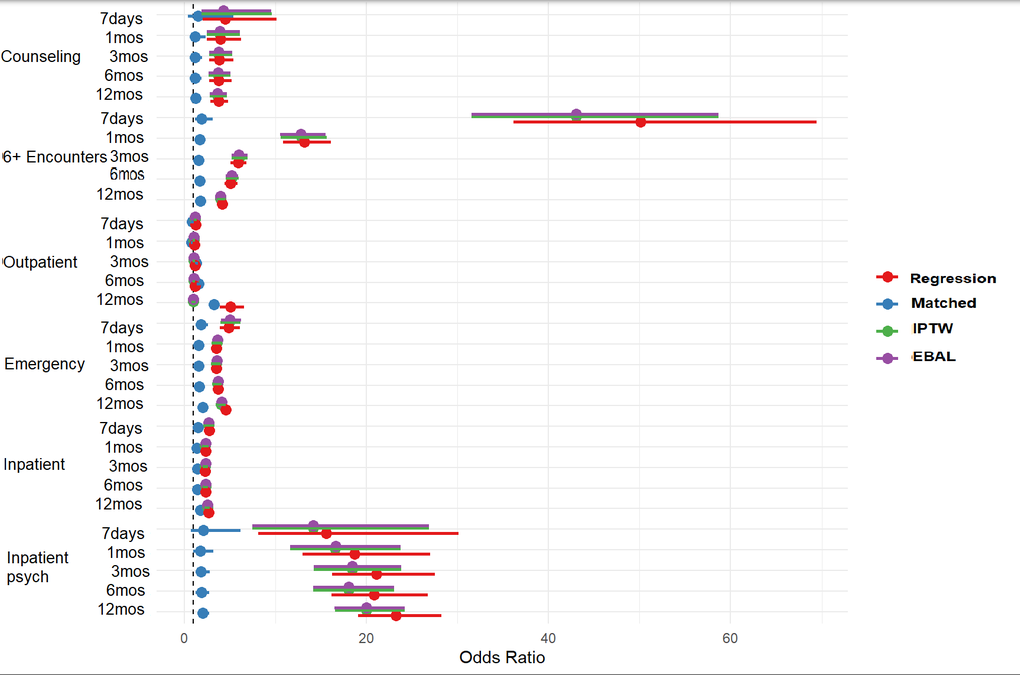

**FIGURE S2.** Effective sample size (ESS), ratio of exposure-to-control size for each estimate and time horizon

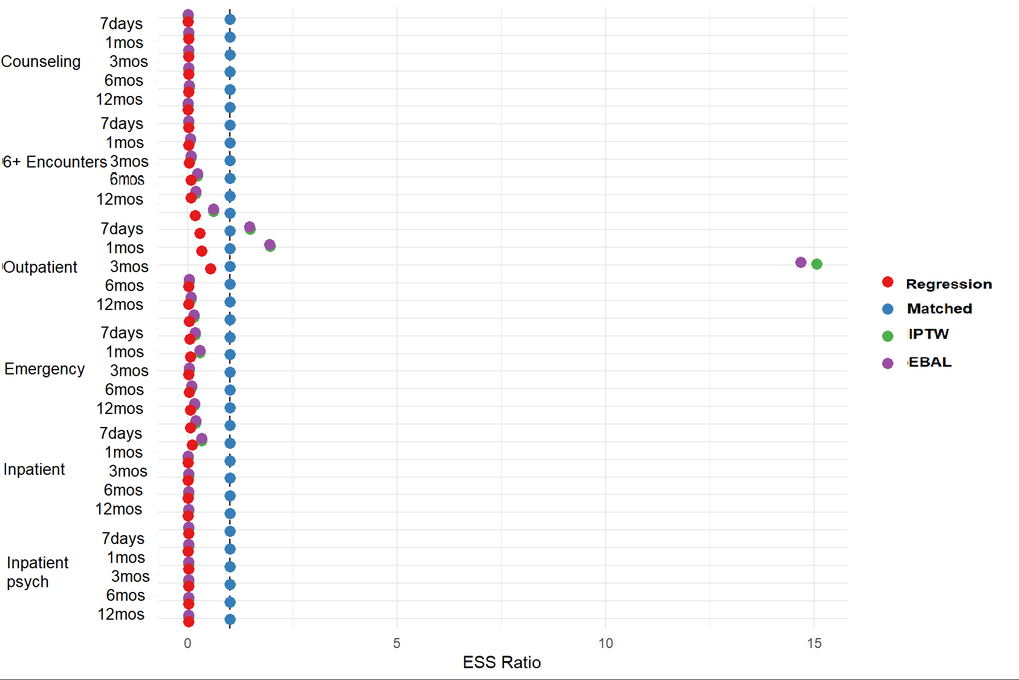
